# Reduction of human Alzheimer’s disease risk and reversal of mouse model cognitive deficit with nucleoside analog use

**DOI:** 10.1101/2023.03.17.23287375

**Authors:** Joseph Magagnoli, Praveen Yerramothu, Kameshwari Ambati, Tammy Cummings, Joseph Nguyen, Claire C. Thomas, Shao-bin Wang, Kaitlyn Cheng, Maksud Juraev, Roshni Dholkawala, Ayami Nagasaka, Meenakshi Ambati, Yosuke Nagasaka, Ashley Ban, Vidya L. Ambati, S. Scott Sutton, Bradley D. Gelfand, Jayakrishna Ambati

## Abstract

Innate immune signaling through the NLRP3 inflammasome has been implicated in the pathogenesis of Alzheimer’s disease (AD), the most prevalent form of dementia. We previously demonstrated that nucleoside reverse transcriptase inhibitors (NRTIs), drugs approved to treat HIV and hepatitis B infections, also inhibit inflammasome activation. Here we report that in humans, NRTI exposure was associated with a significantly lower incidence of AD in two of the largest health insurance databases in the United States. Treatment of aged 5xFAD mice (a mouse model of amyloid-β deposition that expresses five mutations found in familial AD) with Kamuvudine-9 (K-9), an NRTI-derivative with enhanced safety profile, reduced Aβ deposition and reversed their cognitive deficit by improving their spatial memory and learning performance to that of young wild-type mice. These findings support the concept that inflammasome inhibition could benefit AD and provide a rationale for prospective clinical testing of NRTIs or K-9 in AD.

## Main

Alzheimer’s disease (AD) is the leading cause of dementia worldwide, imposing significant burdens on families, societies, and global public health^1,2^. Though considerable progress has been made in discerning the pathogenesis of AD, no clinical cure exists. As the global population ages and the prevalence of AD rises, so does demand for quality therapies^1,2^.

Amyloid-beta (Aβ) oligomers and hyperphosphorylated tau fibrils are thought to promote AD by inducing chronic microglial activation and neuroinflammation that leads to neurodegeneration and cognitive decline^3,4^. A critical effector in the pathogenesis of AD is the NLRP3 inflammasome, a multimeric protein complex that responds to aberrant Aβ and tau aggregation by launching a potent inflammatory response characterized by caspase-1 activation, interleukin-1β (IL-1β) release, and neuronal cell death^5–8^. In turn, NLRP3 activation facilitates further deposition of Aβ plaques and tau fibrils, establishing a positive feedback loop that contributes to the development of AD^9–13^.

NLRP3 inflammasome inhibition may protect against AD progression^9,14^. Previously, we discovered that nucleoside reverse transcriptase inhibitors (NRTIs), which are FDA-approved to treat HIV and Hepatitis B infections, also inhibit inflammasome activation independent of their antiretroviral activity^15^. Prior studies by us and others demonstrated that NRTIs block the NLRP3 inflammatory cascade in a variety of disease states, including models of diabetes, diabetic retinopathy, choroidal neovascularization, and atrophic age-related macular degeneration^16–19^. Given the protective benefits of NLRP3 inflammasome inhibition, we hypothesized that patients with a history of NRTI exposure would be less likely to develop AD.

To test this hypothesis, we analyzed health insurance claims for 263,183 individuals across two national databases spanning a maximum of twenty-one years. In addition, we used the 5xFAD mouse model of AD to evaluate the effects of inflammasome inhibition on brain Aβ deposition and spatial cognitive function. We tested whether Kamuvudine-9 (K-9), an NRTI-derivative engineered to avoid undesirable effects on host polymerases while retaining its therapeutic anti-inflammatory properties, thus enhancing its safety profile^15,19–21^, affected Aβ accumulation and cognitive function.

## Results

### NRTI exposure is associated with reduced risk of developing Alzheimer’s disease

We evaluated the association between NRTI exposure and subsequent development of AD in the Veterans Health Administration (VA), one of the largest integrated healthcare systems in the United States, over a 21-year period. In this database, 63,178 patients (Supplementary Table 1) were identified that met study criteria (at least 50 years of age, diagnosis of HIV or Hepatitis B, and no prior diagnosis of AD). Kaplan-Meier survival estimates revealed better survival (higher probability of no AD diagnosis) across study follow-up for individuals exposed to NRTIs (Fig. 1a, log-rank P < 0.001). In the MarketScan database, which encompasses individuals with employer-based health insurance, a total of 200,005 patients over the years 2006 to 2020 met study inclusion criteria (Supplementary Table 2). Kaplan-Meier survival estimates revealed better survival probability throughout study follow-up in NRTI-exposed individuals compared to those unexposed (Fig 1b, log-rank P < 0.001).

**Figure 1.**
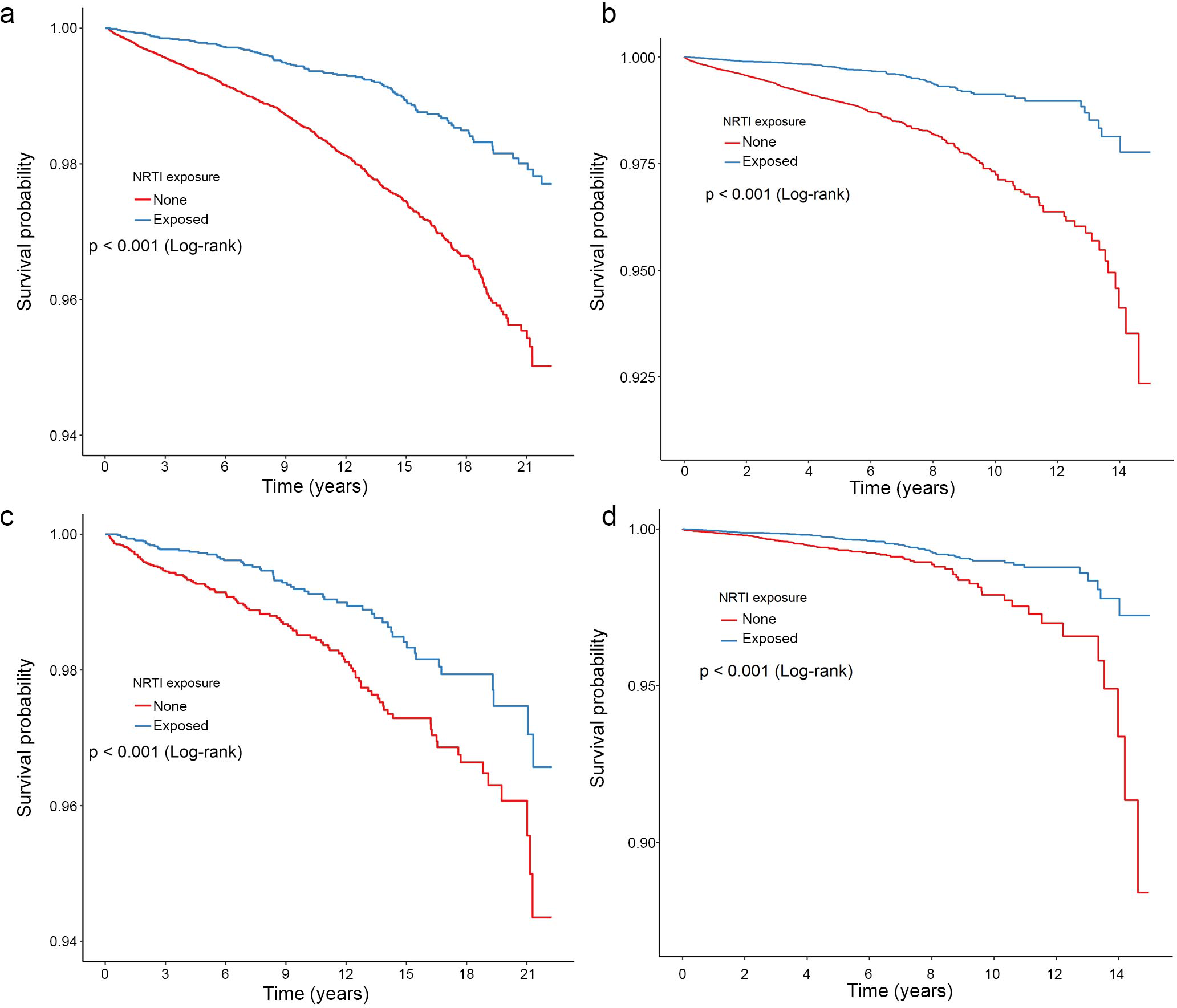
Lower hazard of AD among NRTI users in the VA and Marketscan populations. **a–d**, Survival curves are presented for the VA (**a, c**) and Marketscan (**b, d**) database populations, calculated for each level of nucleoside reverse transcriptase inhibitor (NRTI) exposure in the original (**a, b**) and propensity-score matched (**c, d**) populations. Log-rank P values displayed.

To account for immortal time bias, we also studied NRTI exposure as a time-dependent variable, by calculating the risk of incident AD as a function of cumulative annualized NRTI exposure. Multivariate models were adjusted for important sociodemographic and clinical factors. Demographic data include age at index, race, sex, and smoking status. The Charlson comorbidity score was included as a composite measure of overall health burden. We also adjusted for comorbidities previously reported to be associated with AD (Table 1) including pure hypercholesterolemia, hypertriglyceridemia, hyperlipidemia, ischemic heart disease, other heart disease, hypertension, type 2 diabetes, cerebral infarction, atrial fibrillation, hypothyroidism, hyperthyroidism, depression, traumatic brain injury, alcohol dependence, Parkinson’s disease, generalized anxiety disorder and chronic kidney disease.

**Table 1.** Hazard of AD with NRTI time-dependent exposure, Cox models in original and propensity score (PS)-matched populations.

|  | Original group<br>HR (95% CI) | PS matched<br>HR (95% CI) |
| --- | --- | --- |
| VA Time-dependent exposure:<br>NRTI use per year | **0.915** (0.89, 0.94) | **0.907** (0.862, 0.955) |
| Marketscan Time-dependent exposure:<br>NRTI use per year | **0.894** (0.845, 0.947) | **0.877** (0.821, 0.936) |
| VA Competing Risk of Mortality:<br>NRTI use per year | **0.456** (0.371, 0.56) | **0.514** (0.374, 0.706) |
| **Model covariates** |  |  |
| Age | Race | Sex |
| Charlson comorbidity index | Body mass index | Smoker |
| Pure hypercholesterolemia | Hypertriglyceridemia | Hyperlipidemia |
| Ischemic heart disease | Other heart disease | Hypertension |
| Type 2 diabetes mellitus | Cerebral infarction | Atrial fibrillation |
| Hypothyroidism | Hyperthyroidism | Depression |
| Traumatic Brain Injury | Alcohol dependence | Parkinson's disease |
| Generalized anxiety disorder | Chronic kidney disease |  |

Hazard ratios based on a Cox proportional-hazards model for the original and propensity score (PS)-matched populations in the VA and Marketscan databases, adjusted for the confounding variables listed under Model covariates. Hazard ratios for covariates presented in Supplementary Tables 3 and 4.

In both databases, each additional year of NRTI exposure was associated with reduced hazard of developing AD (Table 1, Supplementary Tables 3 and 4). In the VA sample, each additional year of NRTI exposure was associated with 8.5% reduced hazard of AD (adjusted hazard ratio (aHR), 0.915; 95% CI, 0.89 to 0.94; Supplementary Table 3). In the MarketScan database, there was 10.6% reduced hazard of AD with each additional year of NRTI exposure (aHR, 0.894; 95% CI, 0.845 to 0.947; Supplementary Table 4).

### Propensity score matching analysis

Given that NRTI treatment was not randomized, we used propensity score matching to minimize potential selection bias and differences in baseline characteristics. This process yielded patient cohorts with similar baseline demographic and clinical characteristics. The standardized difference was used to quantify similarities between cohorts. In the VA sample, each cohort contained 8,214 patients and all standardized differences were less than 0.1, indicating excellent balance between cohorts (Supplementary Table 5). In the MarketScan data, each cohort contained 42,297 patients, with all standardized differences less than 0.2 and most less than 0.1 (Supplementary Table 6). As with the original unmatched group results, Kaplan-Meier plots of propensity score-matched populations revealed protective estimates for those exposed to NRTIs compared to those unexposed (log-rank P < 0.001 for both databases; Fig. 1c, Fig. 1d).

We also adjusted for all of the sociodemographic factors, overall health, and comorbidities known to alter risk of AD development that were employed for the original unmatched group analyses in order to control for any residual covariate imbalance. After propensity score matching and adjustment for potential confounders, time-dependent Cox models in each database revealed that each additional year of NRTI exposure was associated with reduced hazard of developing AD (Table 1) (VA: 9.3% reduced hazard per year of NRTI use; aHR, 0.907; 95% CI, 0.862 to 0.955; Supplementary Table 3; MarketScan: 12.3% reduced hazard per year of NRTI exposure; aHR, 0.877; 95% CI, 0.821 to 0.936; Supplementary Table 4). The small differences in hazard ratio estimates between unmatched and propensity score-matched analyses suggest that residual bias in the unmatched analyses was likely minimal.

### Competing Risk of Mortality Analysis

In chronic diseases such as AD, mortality can preclude the diagnosis of AD. Thus, we performed a competing risk regression analysis, which was possible in the VA cohort as the VA contains mortality records. We estimated a Fine and Gray model to account for the competing risk of death. In this analysis, data were organized cross-sectionally, and NRTI exposure was indicated if there was any NRTI use over the study period. After adjusting for potential confounding variables and the competing risk of death, NRTI exposure was associated with 54.4% reduced hazard of developing AD (sub-distribution aHR, 0.456; 95% CI, 0.371 to 0.560; Table 1 and Supplementary Table 7). We also performed the competing risk of mortality analysis in the propensity score-matched population. After adjusting for potential confounding variables and the competing risk of death, NRTI exposure was associated with 48.6% reduced hazard of developing AD (sub-distribution aHR, 0.514; 95% CI, 0.374 to 0.706; Table 1 and Supplementary Table 7). These protective findings, similar to risk reduction observed in the primary analysis, suggest that the differential mortality rates are not responsible for the observed risk reduction of incident AD among NRTI users in the VA cohort.

### K-9 reduced Aβ deposition and restored cognitive function in a mouse model of AD

Since NRTI use was associated with a reduced risk of AD, we sought to determine whether inflammasome inhibition would be beneficial in 5xFAD mice, a translational animal model of AD, by assessing Aβ deposition in the brain as well as spatial memory formation and learning. We treated 24-week-old 5xFAD mice with intraperitoneal administration of K-9 or PBS vehicle for 12 weeks. We found that K-9-treated 5xFAD mice had less Aβ deposition, as monitored by immunofluorescence, in the frontal cortex (Fig. 2a), motor cortex (Fig. 2b), and hippocampus (Fig. 2c), compared to PBS-treated animals.

**Figure 2.**
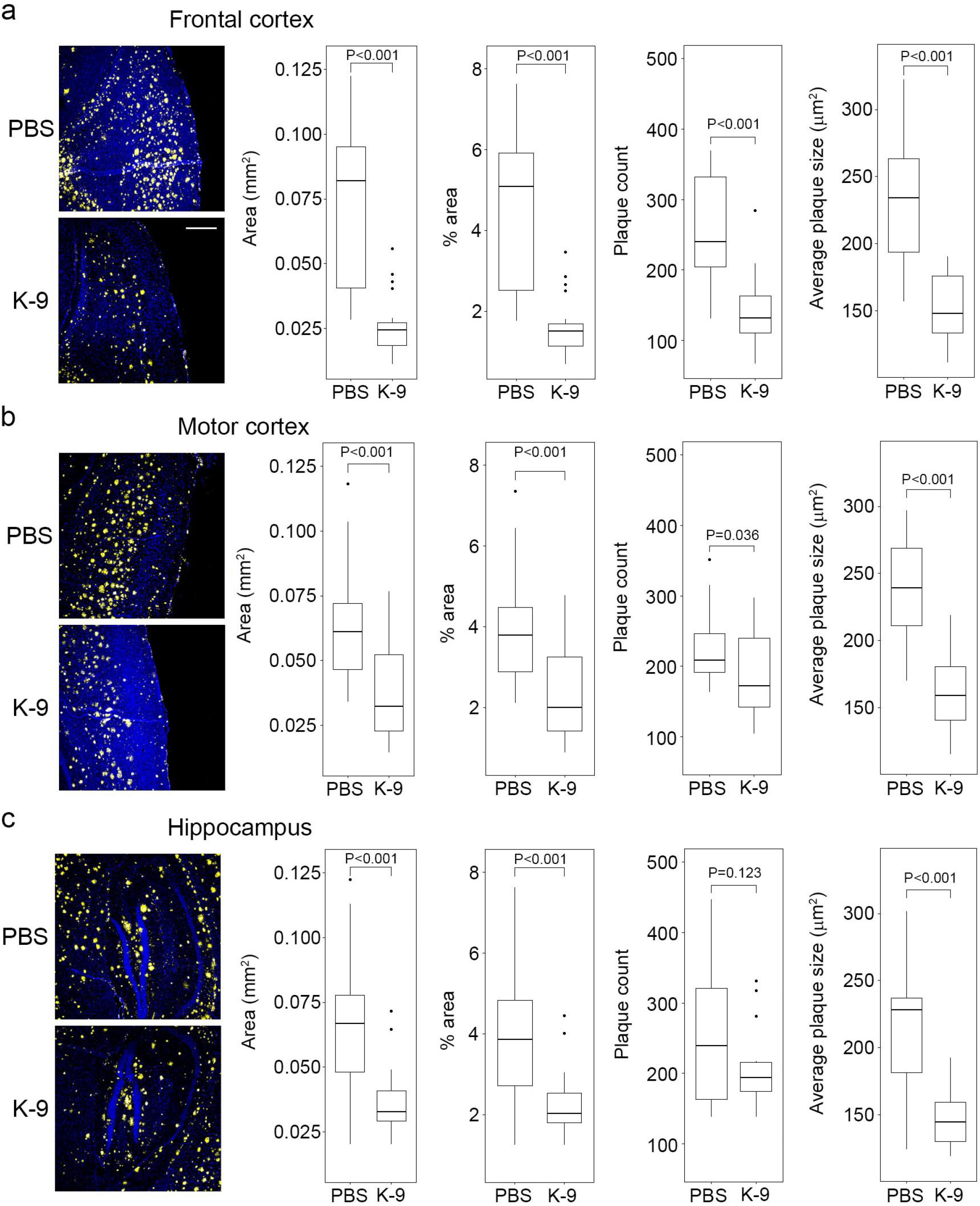
Kamuvudine-9 treatment leads to decreased amyloid-β deposition in 5xFAD mice. **a–c**, Amyloid-β deposition quantification in the brains of 36-week-old 5xFAD mice treated twice daily from 24 weeks of age with Kamuvudine-9 (K-9, i.p. 60 mg/kg twice daily, n = 5) or PBS (i.p. equal volume, n = 5). Amyloid-β quantification performed in five consecutive sections per animal. Left-most panels **(a–c)** show representative images of amyloid-β deposition (yellow) in the frontal cortex, motor cortex and hippocampus. Nuclei stained with DAPI (blue). Scale bar, 200 μm. Box plots (median, interquartile range (IQR), and outliers defined as greater than 1.5 IQRs from the first or third quartiles shown as dots) show quantification of surface area of amyloid-β (Aβ) plaques, % area, number of plaques, and size of plaques. P values from comparisons conducted using mixed effect models fit to account for repeated samples from each brain region within each mouse. Mixed effect models were fit with nested random effects. A random effect for each mouse and a random effect for each brain region within each mouse were included. Fixed effects include the treatment type (K-9, PBS) as well as the brain region.

We assessed spatial memory formation and learning using the Morris water-maze test. We initiated treatment with K-9 or PBS in 5xFAD mice at 24 weeks of age, a time point when this cognitive function in this mouse model is already impaired, as evidenced by their deficits in spatial memory formation and learning compared to 13-week-old 5xFAD mice (P = 0.013, mixed-effects model; Fig. 3, Supplementary Table 8). Following 12 weeks of treatment with K-9, 36-week-old 5xFAD mice exhibited superior spatial memory and learning performance compared to 36-week-old 5xFAD mice treated with PBS (P < 0.001, mixed-effects model contrast estimate; Fig. 3, Supplementary Table 9). Strikingly, 36-week-old K-9-treated 5xFAD mice performed better than when they were 24-weeks-old, and they performed as well as young 18-week-old background strain wild-type mice (P = 0.85, mixed-effects model contrast estimate; Fig. 3, Supplementary Table 9), demonstrating that K-9 treatment not only prevented worsening but induced dramatic improvement in this measure of cognitive function by reversing spatial memory deficits.

**Figure 3.**
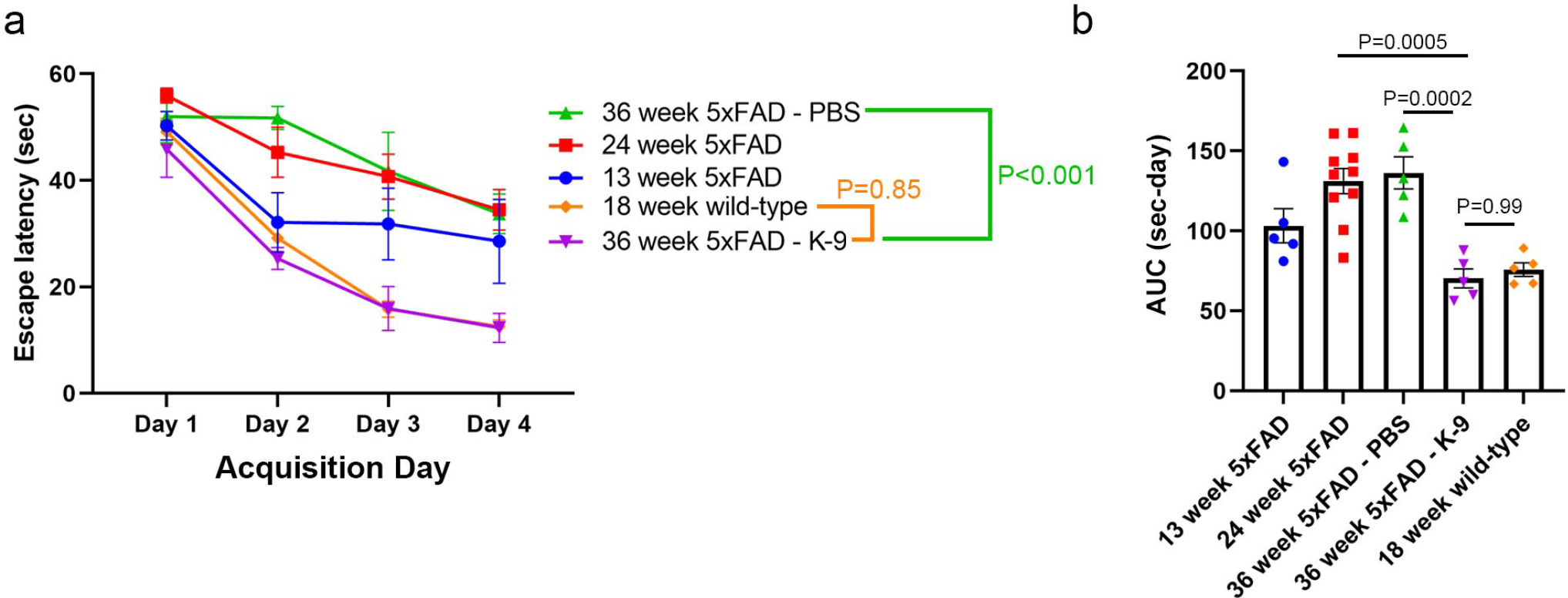
Protective effect of Kamuvudine-9 on spatial memory in 5xFAD mice. **a, b**, Spatial memory was assessed by Morris water maze testing. Time needed to reach the hidden platform (latency) (**a**) and integrated distance (area under the curve, AUC, **b**). **a**, Latency times (mean ± s.e.m.) for each of the acquisition days for 13-week-old 5xFAD mice (n = 5), 24-week-old 5xFAD mice (n = 10), 36-week-old 5xFAD mice treated twice daily from 24 weeks of age with Kamuvudine-9 (K-9, i.p. 60 mg/kg twice daily, n = 5) or PBS (i.p. equal volume, n = 5), and 18-week-old wild-type (n = 5) mice. Comparisons conducted using a mixed effect model with group and day fixed effects along with a random effect for each mouse (Supplementary Table 8). Model based contrasts were estimated comparing the K-9 mice to PBS-treated mice and K-9-treated to wild-type (WT) mice (Supplementary Table 9). Contrast P-values were adjusted using the Bonferroni correction. **b**, AUC using trapezoidal integration for each of the mouse groups (mean ± s.e.m., one-way ANOVA, P values by Tukey’s test).

## Discussion

Innate immune signaling through the NLRP3 inflammasome has been implicated in the pathogenesis of AD^5–13^. A therapy that inhibits inflammasome activation could thus be neuroprotective and improve clinical outcomes in AD. In the present study, we demonstrate that exposure to inflammasome-inhibiting NRTIs^15–19^ is associated with significantly lower incidence of AD in two of the largest health insurance databases in the United States. We also present evidence that K-9 treatment of aged 5xFAD mice dramatically reverses deficits in spatial memory and learning and improves their performance to that of young wild-type mice.

Previously, we demonstrated that K-9 protects against Aβ-induced retinal pigmented epithelium degeneration in a mouse model of age-related macular degeneration^21^. In the present study, we found that K-9 not only reduces Aβ burden in the brains of 5xFAD mice but that it also reverses the impairment in spatial memory formation and learning. Collectively these findings suggest that NLRP3 inflammasome inhibition operates both upstream and downstream of Aβ in the pathogenesis of AD.

Throughout our health insurance database analyses, steps were taken to mitigate potential biases and promote internal validity. At baseline, we adjusted for an extensive number of demographic and clinical variables by including them as fixed risk factor covariates. Given that NRTI treatment was not randomized, we also used propensity score matching to minimize potential selection bias and differences in baseline characteristics. To account for immortal time bias, we studied NRTI exposure as both a binary and a cumulative, time-dependent variable. We also estimated a Fine and Gray model in the VA sample to account for the competing risk of death. Altogether, these measures support the validity and conclusions of database analyses.

Though database samples were limited to patients with HIV or Hepatitis B, it is tempting to speculate that the study results can be generalized to other patient populations. Indeed, K-9 reversed cognitive deficits in 5xFAD mice without these infections. Thus the neuroprotective effects of NRTIs and K-9 in AD likely extend beyond the setting of coexisting viral infection.

This is consistent with prior studies by us and others that demonstrate the anti-inflammatory benefits of NRTIs and K-9 in other models of noninfectious diseases^16–19^. Nonetheless, prospective randomized controlled trials in humans are warranted to gain better insight into the effects of NRTIs or K-9 on clinical outcomes in AD. Indeed, there are ongoing trials of two NRTIs, lamivudine (NCT04552795) and emtricitabine (NCT04500847), in AD. It bears noting that isolated NRTI use can facilitate the development of viral resistance^22^. Further, NRTIs have been associated with off-target mitochondrial toxicity due to their inhibition of mitochondrial gamma polymerase^23^. Thus it merits testing whether K-9, which lacks these drawbacks^15^ could be a safer alternative to NRTIs in the clinical setting.

As the prevalence of AD rises, so does demand for quality, disease-modifying therapies^1,2^. The repurposing of existing, approved therapies could accelerate drug development. Though not all NRTIs may be suitable candidates (e.g., first generation stavudine and didanosine with increased off-target effects), newer NRTIs and their derivatives may be optimal for repurposing. Our data suggest that K-9, an NRTI-derivative with enhanced safety profile^15,20^, could provide dramatic neuroprotection, and provide a rationale for clinical testing of NRTIs or K-9 in AD.

## Online Methods

### Data sources

Data were evaluated from two health insurance claims databases: The United States Veterans Health Administration database (which includes healthcare claims from the VA Informatics and Computing Infrastructure for approximately 11 million individuals) for the years 2000–2021, and IBM MarketScan (which includes employer-based health insurance claims of approximately 158 million individuals) for the years 2006–2020. Codes from the International Classification of Diseases, 9^th^ and 10^th^ Revisions, Clinical Modification (ICD-9-CM and ICD-10-CM) were used to identify individuals with diagnoses of interest. This drug disease cohort study was conducted using data from the U.S. Department of Veterans Affairs. The VA Informatics and Computing Infrastructure (VINCI) was utilized to obtain individual-level information on demographics, administrative claims and pharmacy dispensation. The study was conducted in compliance with the Department of Veterans Affairs requirements and received Institutional Review Board (IRB) and Research and Development approval. All data within the Marketscan database are Health Insurance Portability and Accountability Act-compliant and thus were deemed exempt from IRB approval by the University of Virginia IRB.

### Study population

Patients were included in analyses if they had a medical claim for HIV/AIDS or hepatitis B (HBV) during the study period and were at least 50-years-old at the study index (the first date of HIV/HBV per database). Patients that had a prior diagnosis of AD were excluded. Eligible patients were followed until death (VA), incident AD (VA and Marketscan), or end of eligibility (MarketScan). The date of death was determined by the VA vital status file, which was sourced from the Social Security Administration Death Master File and Veterans Benefits Administration (VBA) Beneficiary Identification Records Locator Subsystem Death File.

### Exposure and outcome definitions

Individuals were classified as receiving NRTIs (i.e., lamivudine, zidovudine, abacavir, didanosine, stavudine, tenofovir, emtricitabine, entecavir, adefovir, telbivudine, entecavir), if at least one pharmacy prescription for these medications was filled. Pharmacy records were filtered by either text search (VA) or by national drug code (MarketScan database). The primary outcome was incident AD.

### Health insurance database statistical analyses

The key predictor was NRTI exposure. Cox regression was used to estimate the hazard of developing AD in relation to NRTI exposure, with adjustment for baseline covariates including age, race, (available for the VA), sex, Charlson comorbidity score, pure hypercholesterolemia, hypertriglyceridemia, hyperlipidemia, ischemic heart disease, other heart disease, hypertension, type II diabetes mellitus, cerebral infarction, atrial fibrillation, hypothyroidism, hyperthyroidism, depression, traumatic brain injury, alcohol dependence, Parkinson’s disease, generalized anxiety disorder, chronic kidney disease), and 95% confidence intervals were constructed for hazard ratios based on standard errors derived from the model. All covariates were flagged during the period between 1 year prior to the study index and the study index date. T-tests and chi-square tests were used to compare continuous variables (age, Charlson comorbidity index) and categorical variables (race, sex), respectively. Given that death would preclude a diagnosis of AD, a Fine and Gray competing risk model was fit to the VA sample, with corresponding sub-distribution hazard ratios and 95% confidence intervals. Given that NRTI treatment was not randomized, we also used propensity score (PS) matching (accounting for both the demographic and clinical factors above) to minimize potential selection bias. To create the matched cohorts, we used greedy nearest neighbor 1:1 PS matching. Kaplan-Meier survival curves are reported for both the original and matched cohorts.

### Animals

All experiments involving animals were approved by the University of Virginia Institutional Animal Care and Use Committee (IACUC). 5xFAD transgenic mice on the B6SJLF1/J background (MMRRC Strain #034840-JAX) and wild-type B6SJLF1/J mice (Strain #100012) were purchased from The Jackson Laboratory (Bar Harbor, ME, USA). All mice were housed under controlled conditions (12-hour light/dark cycle, 5 mice housed per cage, and standard chow diet).

### Drug treatments

At 24 weeks of age, 5xFAD mice were randomized into one of two groups (5 mice/group) to receive intraperitoneal (i.p.) injections of either K-9 (trimethyl-3TC, 60 mg/kg dissolved in 1x PBS) or a matched volume of 1x PBS, twice daily for 12 weeks. K-9 was synthesized as previously described^21,24^. Injections were administered in masked fashion.

### Morris Water Maze testing

To evaluate spatial memory, the Morris Water Maze (MWM) was performed. Mice were housed in the testing room at least 1 hour prior to starting. Testing was performed between 1300 and 1600 each day, and researchers were masked to genotype and treatment information. A hidden platform (10 cm diameter, 1 cm below the surface) was located in a water maze pool (1 m diameter) filled with opaque water (22–23°C). The pool was divided into four quadrants, with signs posted (triangle, circle, square, and X) to orient the mice. In total, mice were evaluated across four consecutive training days (4 trials/day, each separated by at least 30 min). During the four training days, mice were trained to locate the hidden platform. Escape latency was noted as the time taken to locate the hidden platform. Mean latency was calculated as the four-trial daily average. If a mouse failed to find the platform within 60 sec, it was manually placed on the platform, where it was allowed to remain for 120 sec after the first trial to observe its surroundings (allowed 10 sec after any subsequent trial). Data were recorded and analyzed using an automated video tracking system EthoVision XT 13 (Noldus, Wageningen, the Netherlands).

Comparison between K-9- and PBS-treated mice was conducted using a mixed effect model with group and day fixed effects along with a random effect for each mouse. Model based contrasts were estimated comparing the K-9 mice to PBS-treated mice and K-9-treated mice to wild-type (WT) mice. Contrast P-values were adjusted using the Bonferroni correction. Mixed effect models were fit with the R package LME4 and model-based P-values extracted using the R package lmerTest. Contrasts were estimated using the R package multcomp. Areas under the curve (AUCs) were calculated using trapezoidal integration. Groups were compared using one-way analysis of variance (ANOVA) with post hoc Tukey’s test.

### Immunofluorescence of Aβ deposition

Immunofluorescence staining was performed to quantify the deposition of brain Aβ in 5XFAD mice. Mice were perfused with cold saline, followed by paraformaldehyde (PFA). Brain sections (sagittal, 7 μm) were collected from the PFA-fixed hemibrains (4% PFA overnight). Sections were blocked in donkey blocking buffer (2% normal donkey serum, 1% BSA, 0.1% Triton-x in 1xPBS) for 1 hour at 37°C, stained with primary antibody overnight at 4°C (1:500, rabbit anti-Aβ, Abcam, ab201061), and conjugated with secondary antibody (1:1000, donkey anti-rabbit Alexa 555, Invitrogen) for 1 hour at room temperature. All sections were counter-stained with DAPI (P36935, ThermoFisher) and imaged using a Nikon A1R laser confocal microscope.

Aβ quantification was performed in masked fashion using ImageJ (version 1.53q). Images were normalized to anatomic boundaries, and plaques were identified via automatic thresholding. Images were converted to an 8-bit binary format and “fill holes” and “watershed” algorithms were applied. Plaque number, area, and average size were calculated using the ‘analyze particles’ plugin of ImageJ (NIH). Aβ area fraction was calculated by dividing the total plaque area by the area of the microscopic field. Five serial sections per hemibrain per mouse were averaged to obtain the reported results.

To compare K-9-treated vs. PBS-treated mice in terms of Aβ quantification, mixed effect models were fit to account for repeated samples from each brain region within each mouse. Mixed effect models were fit with nested random effects using the R package LME4. A random effect for each mouse and a random effect for each brain region within each mouse were included. Fixed effects include the treatment type (K-9, PBS) as well as the brain region. P-values for fixed effects were extracted with the lmerTest package.

## Data Availability

All data produced in the present work are contained in the manuscript

## Acknowledgments

We thank J. Thanos, L.I. Baldridge, J. Hu, and S. Staples for technical assistance. J.A. discloses support from the UVA Strategic Investment Fund and National Institutes of Health (NIH) grants (R01EY028027, R01EY029799, R01EY031039, R01AG082108), the DuPont Guerry, III, Professorship, and a gift from Mr. and Mrs. Eli W. Tullis. B.D.G. discloses support from NIH grants (R01EY028027, R01EY031039, R01AG082108, R01EY032512). S.S.S., J.M., and T.H.C. disclose support from NIH grant R01DA054992 and the South Carolina Center for Rural and Primary Healthcare for projects unrelated to this study. This paper represents original research conducted using data from the Department of Veterans Affairs. This material is the result of work supported with resources and the use of facilities at the Dorn Research Institute, Columbia VA Health Care System, Columbia, South Carolina. The content of this article is solely the responsibility of the authors and does not necessarily represent the official views of the US Department of Veterans Affairs, nor does mention of trade names, commercial products or organizations imply endorsement by the US government.

## Author contributions

J.M., P.Y., K.A., T.C., J.N., S.W., K.C., M.J., R.D., A.N., M.A., Y.N., A.B., V.L.A., S.S.S., B.D.G., and J.A. performed experiments or evaluated and interpreted the data. J.A., J.M., and S.S.S. conceived the study concept and design. J.M., C.T., B.D.G, and J.A. wrote the manuscript. All authors took part in the interpretation of the results, commented on the manuscript, and had final responsibility for the decision to submit for publication.

