## Supplemental Tables for "Reduction of human Alzheimer’s disease risk and reversal of mouse model cognitive deficit with nucleoside analog use"

**Supplementary Table 1. VA Original group baseline characteristics.**

| | Variable | NRTI unexposed N=43493 | NRTI exposed N=19685 | P value |
|---|---|---|---|---|
| HIV | | 6674(15.345%) | 14864(75.509%) | <0.001 |
| HBV | | 37200(85.531%) | 6886(34.981%) | <0.001 |
| Age | | 61.77(9.713) | 59.9(9.201) | <0.001 |
| Race | Black | 12376(28.455%) | 7928(40.274%) | <0.001 |
| | Other/unknown | 5661(13.016%) | 2173(11.039%) | |
| | White | 25456(58.529%) | 9584(48.687%) | |
| Sex | Female | 1189(2.734%) | 349(1.773%) | <0.001 |
| | Male | 42304(97.266%) | 19336(98.227%) | |
| Charlson comorbidity | | 1.52(2.081) | 0.75(1.598) | <0.001 |
| Smoker | | 18147(41.724%) | 8193(41.621%) | 0.814 |
| BMI | <18.5 | 1028(2.364%) | 746(3.79%) | <0.001 |
| | 18.5-24.9 | 12008(27.609%) | 7932(40.295%) | |
| | 25-29.9 | 16006(36.801%) | 6643(33.747%) | |
| | 30+ | 13846(31.835%) | 4074(20.696%) | |
| | Missing | 605(1.391%) | 290(1.473%) | |
| Pure hypercholesterolemia | | 2249(5.171%) | 381(1.935%) | <0.001 |
| Hypertriglyceridemia | | 2126(4.888%) | 510(2.591%) | <0.001 |
| Hyperlipidemia | | 10760(24.74%) | 2433(12.36%) | <0.001 |
| Ischemic heart disease | | 6124(14.08%) | 1256(6.38%) | <0.001 |
| Other Heart disease | | 5688(13.078%) | 1043(5.298%) | <0.001 |
| Hypertension | | 16616(38.204%) | 3597(18.273%) | <0.001 |
| T2DM | | 7428(17.079%) | 1543(7.838%) | <0.001 |
| Cerebral infarction | | 262(0.602%) | 51(0.259%) | <0.001 |
| Atrial fibrillation | | 1104(2.538%) | 140(0.711%) | <0.001 |
| Hypothyroidism | | 1244(2.86%) | 193(0.98%) | <0.001 |
| Hyperthyroidism | | 133(0.306%) | 28(0.142%) | <0.001 |
| Depression | | 7662(17.617%) | 1547(7.859%) | <0.001 |
| TBI | | 195(0.448%) | 41(0.208%) | <0.001 |
| Alcohol dependence | | 5772(13.271%) | 924(4.694%) | <0.001 |
| Parkinson's disease | | 170(0.391%) | 16(0.081%) | <0.001 |
| Generalized anxiety disorder | | 653(1.501%) | 137(0.696%) | <0.001 |
| Chronic kidney disease | | 1735(3.989%) | 360(1.829%) | <0.001 |
| Year of HIV/HBV dx | | 2008.62(6.327) | 2008.09(6.71) | <0.001 |
| NRTI use (years) | | 0(0) | 5.8(5.1) | <0.001 |

HIV, Human immunodeficiency virus. HBV, Hepatitis B virus. BMI, body mass index. T2DM, type 2 diabetes mellitus. TBI, traumatic brain injury. NRTI, nucleoside reverse transcriptase inhibitor.

**Supplementary Table 2. Marketscan Original group baseline characteristics.**

| | | NRTI unexposed N=136019 | NRTI exposed N=63986 | P value | Standardized difference |
|---|---|---|---|---|---|
| HIV | | 44340(32.84%) | 50918(79.577%) | <0.001 | 1.07 |
| HBV | | 92534(68.534%) | 15786(24.671%) | <0.001 | 0.977 |
| Age | | 59.36(7.23) | 56.92(5.1) | <0.001 | 0.39 |
| Sex | Male | 80013(59.261%) | 51424(80.368%) | <0.001 | 0.472 |
| | Female | 55006(40.739%) | 12562(19.632%) | | 0.472 |
| Charlson comorbidity | | 1.23(2.201) | 0.37(1.166) | <0.001 | 0.486 |
| Smoker | | 8316(6.159%) | 1164(1.819%) | <0.001 | 0.226 |
| Obese | | 7743(5.735%) | 912(1.425%) | <0.001 | 0.234 |
| Pure hypercholesterolemia | | 12808(9.486%) | 1483(2.318%) | <0.001 | 0.309 |
| Hypertriglyceridemia | | 9761(7.229%) | 1445(2.258%) | <0.001 | 0.232 |
| Hyperlipidemia | | 26873(19.903%) | 3852(6.02%) | <0.001 | 0.423 |
| Ischemic heart disease | | 12397(9.182%) | 1697(2.652%) | <0.001 | 0.277 |
| Other Heart disease | | 17521(12.977%) | 1958(3.06%) | <0.001 | 0.37 |
| Hypertension | | 47902(35.478%) | 7751(12.114%) | <0.001 | 0.571 |
| T2DM | | 23601(17.48%) | 3831(5.987%) | <0.001 | 0.363 |
| Cerebral infarction | | 1626(1.204%) | 173(0.27%) | <0.001 | 0.104 |
| Atrial fibrillation | | 4276(3.167%) | 411(0.642%) | <0.001 | 0.191 |
| Hypothyroidism | | 11141(8.251%) | 1010(1.578%) | <0.001 | 0.313 |
| Hyperthyroidism | | 1244(0.921%) | 146(0.228%) | <0.001 | 0.095 |
| Depression | | 8221(6.089%) | 1726(2.697%) | <0.001 | 0.166 |
| TBI | | 596(0.441%) | 68(0.106%) | <0.001 | 0.06 |
| Alcohol dependence | | 1332(0.987%) | 177(0.277%) | <0.001 | 0.087 |
| Parkinson’s disease | | 372(0.276%) | 44(0.069%) | <0.001 | 0.045 |
| Generalized anxiety disorder | | 3592(2.66%) | 905(1.414%) | <0.001 | 0.092 |
| Chronic kidney disease | | 9115(6.751%) | 1127(1.761%) | <0.001 | 0.248 |
| Year of HIV/HBV dx | | 2011.99(3.788) | 2012.79(4.301) | <0.001 | 0.196 |
| NRTI duration (years) | | 0(0) | 2.4(2.307) | <0.001 | 1.470 |

HIV, Human immunodeficiency virus. HBV, Hepatitis B virus. BMI, body mass index. T2DM, type 2 diabetes mellitus. TBI, traumatic brain injury. NRTI, nucleoside reverse transcriptase inhibitor.

**Supplementary Table 3. VA Cox models, hazard of AD: Time-dependent NRTI exposure**

| | | Original group<br>HR(95% CI) | PS matched<br>HR(95% CI) |
|---|---|---|---|
| NRTI use per year | | 0.915 (0.89-0.94) | 0.907 (0.862-0.955) |
| Age | | 1.027 (1.025-1.03) | 1.033 (1.027-1.038) |
| Race | Black vs. white | 0.934 (0.793-1.099) | 0.952 (0.679-1.335) |
| | Other/Unknown vs. white | 0.912 (0.717-1.159) | 1.324 (0.861-2.036) |
| Sex | Female vs. male | 0.445 (0.23-0.862) | 0.542 (0.132-2.227) |
| Charlson comorbidity | | 1.184 (1.137-1.234) | 1.141 (1.043-1.248) |
| BMI | 18.5-24.9 vs. missing | 1 (0.513-1.952) | 3.143 (0.433-22.82) |
| | 25-29.9 vs. missing | 0.83 (0.426-1.614) | 2.434 (0.335-17.675) |
| | 30+ vs. missing | 0.63 (0.322-1.235) | 1.441 (0.195-10.641) |
| | <18.5 vs. missing | 1.039 (0.448-2.409) | 2.522 (0.26-24.488) |
| Smoker | | 0.427 (0.361-0.505) | 0.426 (0.295-0.614) |
| Pure hypercholesterolemia | | 1.245 (0.958-1.618) | 1.14 (0.583-2.228) |
| Hypertriglyceridemia | | 0.913 (0.637-1.307) | 1.208 (0.56-2.606) |
| Hyperlipidemia | | 0.916 (0.763-1.099) | 0.892 (0.595-1.335) |
| Ischemic heart disease | | 1.092 (0.875-1.365) | 1.142 (0.694-1.88) |
| Other Heart disease | | 1.217 (0.946-1.565) | 1.682 (0.993-2.851) |
| Hypertension | | 1.21 (1.027-1.426) | 1.378 (0.976-1.945) |
| T2DM | | 1.097 (0.894-1.345) | 1.081 (0.686-1.703) |
| Cerebral infarction | | 1.955 (1.035-3.693) | 7.49 (2.662-21.073) |
| Atrial fibrillation | | 1.232 (0.811-1.872) | 0.359 (0.072-1.784) |
| Hypothyroidism | | 1.205 (0.825-1.761) | 1.391 (0.55-3.52) |
| Hyperthyroidism | | 0.953 (0.303-2.992) | 1.955 (0.251-15.222) |
| Depression | | 1.099 (0.898-1.346) | 0.999 (0.612-1.632) |
| TBI | | 1.805 (0.746-4.367) | - |
| Alcohol dependence | | 0.87 (0.665-1.139) | 0.707 (0.333-1.502) |
| Parkinson's disease | | 5.419 (3.172-9.257) | - |
| Generalized anxiety disorder | | 1.207 (0.718-2.029) | - |
| Chronic kidney disease | | 0.599 (0.395-0.909) | 0.566 (0.218-1.473) |

HIV, Human immunodeficiency virus. HBV, Hepatitis B virus. BMI, body mass index. T2DM, type 2 diabetes mellitus. TBI, traumatic brain injury. NRTI, nucleoside reverse transcriptase inhibitor.

**Supplementary Table 4. Marketscan Cox models, hazard of AD: Time-dependent NRTI exposure**

| | Original group HR (95% CI) | PS matched HR (95% CI) |
|---|---|---|
| NRTI use per year | 0.894 (0.845-0.947) | 0.877 (0.821-0.936) |
| Age | 1.145 (1.138-1.152) | 1.163 (1.149-1.177) |
| Sex Male vs. Female | 0.876 (0.764-1.005) | 0.897 (0.689-1.169) |
| Smoker | 1.188 (0.902-1.563) | 1.995 (1.063-3.746) |
| Obesity | 0.956 (0.682-1.34) | 0.962 (0.353-2.621) |
| Charlson comorbidity | 1.047 (1.011-1.084) | 1.095 (1.006-1.191) |
| Pure hypercholesterolemia | 0.882 (0.713-1.09) | 1.119 (0.648-1.93) |
| Hypertriglyceridemia | 0.965 (0.736-1.267) | 0.566 (0.224-1.431) |
| Hyperlipidemia | 0.962 (0.812-1.14) | 0.953 (0.639-1.42) |
| Ischemic heart disease | 1.163 (0.964-1.403) | 1.135 (0.748-1.724) |
| Other Heart disease | 1.225 (1.019-1.474) | 0.958 (0.632-1.452) |
| Hypertension | 1.014 (0.855-1.201) | 0.839 (0.6-1.173) |
| T2DM | 1.383 (1.163-1.644) | 1.288 (0.872-1.9) |
| Cerebral infarction | 1.499 (1.066-2.109) | 2.879 (1.197-6.925) |
| Atrial fibrillation | 0.657 (0.505-0.854) | 0.799 (0.352-1.814) |
| Hypothyroidism | 1.036 (0.834-1.287) | 1.177 (0.615-2.254) |
| Hyperthyroidism | 0.876 (0.425-1.803) | 0.886 (0.112-6.994) |
| Depression | 1.709 (1.359-2.149) | 1.685 (0.98-2.897) |
| TBI | 1.396 (0.706-2.761) | 0.674 (0.055-8.199) |
| Alcohol dependence | 1.673 (0.934-2.995) | 0 (0-0) |
| Parkinson's disease | 4.987 (3.4-7.313) | 9.43 (4.904-18.133) |
| Generalized anxiety disorder | 1.21 (0.801-1.828) | 1.079 (0.365-3.189) |
| Chronic kidney disease | 1.06 (0.851-1.321) | 0.778 (0.42-1.44) |
| Year of diagnosis | 0.957 (0.936-0.978) | 0.972 (0.934-1.011) |

HIV, Human immunodeficiency virus. HBV, Hepatitis B virus. BMI, body mass index. T2DM, type 2 diabetes mellitus. TBI, traumatic brain injury. NRTI, nucleoside reverse transcriptase inhibitor.

**Supplementary Table 5. VA: Propensity Score-Matched cohorts baseline characteristics.**

| | Variable | NRTI unexposed N=8214 | NRTI exposed N=8214 | P value | Standardized difference |
|---|---|---|---|---|---|
| HIV | | 3693(44.96%) | 3695(44.98%) | 0.987 | 0 |
| HBV | | 4714(57.39%) | 4693(57.13%) | 0.752 | 0.005 |
| Age | | 61.54(9.21) | 61.91(9.25) | <0.001 | 0.04 |
| Race | Black | 2692(32.77%) | 2851(34.71%) | <0.001 | 0.041 |
| | Other/unknown | 1154(14.05%) | 1310(15.95%) | | 0.053 |
| | White | 4368(53.18%) | 4053(49.34%) | | 0.077 |
| Sex | Female | 178(2.17%) | 175(2.13%) | 0.914 | 0.002 |
| | Male | 8036(97.83%) | 8039(97.87%) | | 0.002 |
| Charlson comorbidity | | 1.12(1.83) | 1.25(1.99) | 0.001 | 0.068 |
| Smoker | | 2992(36.43%) | 2993(36.44%) | 1 | 0 |
| BMI | <18.5 | 227(2.76%) | 303(3.69%) | <0.001 | 0.052 |
| | 18.5-24.9 | 2551(31.06%) | 2755(33.54%) | | 0.053 |
| | 25-29.9 | 2971(36.17%) | 2769(33.71%) | | 0.052 |
| | 30+ | 2327(28.33%) | 2239(27.26%) | | 0.024 |
| | Missing | 138(1.68%) | 148(1.80%) | | 0.009 |
| Pure hypercholesterolemia | | 192(2.34%) | 236(2.87%) | 0.035 | 0.034 |
| Hypertriglyceridemia | | 286(3.48%) | 327(3.98%) | 0.1 | 0.026 |
| Hyperlipidemia | | 1501(18.27%) | 1665(20.27%) | 0.001 | 0.051 |
| Ischemic heart disease | | 727(8.85%) | 822(10.00%) | 0.012 | 0.04 |
| Other Heart disease | | 561(6.83%) | 602(7.33%) | 0.224 | 0.019 |
| Hypertension | | 2099(25.55%) | 2153(26.21%) | 0.345 | 0.015 |
| T2DM | | 916(11.15%) | 937(11.41%) | 0.622 | 0.008 |
| Cerebral infarction | | 31(0.377%) | 28(0.34%) | 0.794 | 0.006 |
| Atrial fibrillation | | 58(0.71%) | 61(0.74%) | 0.854 | 0.004 |
| Hypothyroidism | | 133(1.62%) | 108(1.32%) | 0.119 | 0.025 |
| Hyperthyroidism | | 19(0.23%) | 14(0.17%) | 0.486 | 0.014 |
| Depression | | 831(10.12%) | 866(10.54%) | 0.383 | 0.014 |
| TBI | | 23(0.28%) | 28(0.34%) | 0.575 | 0.011 |
| Alcohol dependence | | 545(6.64%) | 444(5.41%) | 0.001 | 0.052 |
| Parkinson’s disease | | 4(0.049%) | 8(0.097%) | 0.386 | 0.018 |
| Generalized anxiety disorder | | 65(0.79%) | 75(0.91%) | 0.445 | 0.013 |
| Chronic kidney disease | | 196(2.39%) | 213(2.59%) | 0.423 | 0.013 |
| Year of HIV/HBV dx | | 2009.27(6.67) | 2009.65(6.74) | 0.001 | 0.057 |
| NRTI use (year) | | 0(0) | 4.44(4.48) | <0.001 | NA |

HIV, Human immunodeficiency virus. HBV, Hepatitis B virus. BMI, body mass index. T2DM, type 2 diabetes mellitus. TBI, traumatic brain injury. NRTI, nucleoside reverse transcriptase inhibitor.

**Supplementary Table 6. Truven: Propensity Score Matched cohorts baseline characteristics.**

| | | NRTI unexposed N=48297 | NRTI exposed N=48297 | P value | Standardized difference |
|---|---|---|---|---|---|
| HIV | | 35073(72.619%) | 35229(72.942%) | 0.263 | 0.007 |
| HBV | | 14799(30.642%) | 14671(30.377%) | 0.375 | 0.006 |
| Age | | 57.5(5.67) | 56.64(5.446) | <0.001 | 0.155 |
| Sex | Male | 35513(73.53%) | 36497(75.568%) | <0.001 | 0.047 |
| | Female | 12784(26.47%) | 11800(24.432%) | <0.001 | 0.047 |
| Charlson comorbidity | | 0.5(1.295) | 0.49(1.314) | <0.001 | 0.008 |
| Smoker | | 1082(2.24%) | 1158(2.398%) | 0.109 | 0.01 |
| Obese | | 932(1.93%) | 908(1.88%) | 0.588 | 0.004 |
| Pure hypercholesterolemia | | 1541(3.191%) | 1476(3.056%) | 0.236 | 0.008 |
| Hypertriglyceridemia | | 1372(2.841%) | 1415(2.93%) | 0.419 | 0.005 |
| Hyperlipidemia | | 3775(7.816%) | 3816(7.901%) | 0.632 | 0.003 |
| Ischemic heart disease | | 1635(3.385%) | 1651(3.418%) | 0.79 | 0.002 |
| Other Heart disease | | 2014(4.17%) | 1955(4.048%) | 0.347 | 0.006 |
| Hypertension | | 7851(16.256%) | 7581(15.697%) | 0.0182 | 0.015 |
| T2DM | | 3844(7.959%) | 3796(7.86%) | 0.575 | 0.004 |
| Cerebral infarction | | 158(0.327%) | 172(0.356%) | 0.473 | 0.005 |
| Atrial fibrillation | | 422(0.874%) | 407(0.843%) | 0.625 | 0.003 |
| Hypothyroidism | | 1079(2.234%) | 1008(2.087%) | 0.121 | 0.01 |
| Hyperthyroidism | | 133(0.275%) | 146(0.302%) | 0.472 | 0.005 |
| Depression | | 1391(2.88%) | 1641(3.398%) | <0.001 | 0.03 |
| TBI | | 62(0.128%) | 68(0.141%) | 0.661 | 0.004 |
| Alcohol dependence | | 152(0.315%) | 175(0.362%) | 0.223 | 0.008 |
| Parkinson's disease | | 48(0.099%) | 44(0.091%) | 0.754 | 0.003 |
| Generalized anxiety disorder | | 693(1.435%) | 875(1.812%) | <0.001 | 0.03 |
| Chronic kidney disease | | 1106(2.29%) | 1101(2.28%) | 0.931 | 0.001 |
| Year of HIV/HBV dx | | 2011.93(3.924) | 2012.43(4.13) | <0.001 | 0.124 |
| NRTI duration (years) | | 0(0) | 2.43 (2.33) | <0.001 | NA |

HIV, Human immunodeficiency virus. HBV, Hepatitis B virus. BMI, body mass index. T2DM, type 2 diabetes mellitus. TBI, traumatic brain injury. NRTI, nucleoside reverse transcriptase inhibitor.

**Supplementary Table 7. VA Competing risk model, death as a competing risk. Sub-distribution hazard ratios (sHR).**

| | | Original group sHR(95% CI) | PS matched sHR(95% CI) |
|---|---|---|---|
| NRTI use per year | | 0.456 (0.371-0.56) | 0.514 (0.374-0.706) |
| Age | | 1.027 (1.024-1.03) | 1.032 (1.023-1.04) |
| Race | Black vs. white | 1.027 (0.869-1.213) | 0.9712 (0.994-0.706) |
| | Other/Unknown vs. white | 0.692 (0.539-0.888) | 0.8962 (1.03-0.664) |
| Sex | Female vs. white | 0.522 (0.261-1.046) | 0.6246 (0.699-0.167) |
| Charlson comorbidity | | 1.016 (0.975-1.059) | 1.002 (0.918-1.095) |
| BMI | 18.5-24.9 vs. missing | 1.241 (0.631-2.443) | 3.613 (0.497-26.251) |
| | 25-29.9 vs. missing | 1.235 (0.63-2.421) | 3.142 (0.432-22.875) |
| | 30+ vs. missing | 0.979 (0.496-1.929) | 2.213 (0.3-16.329) |
| | <18.5 vs. missing | 0.904 (0.387-2.108) | 1.961 (0.202-19.032) |
| Smoker | | 0.509 (0.43-0.603) | 0.49 (0.333-0.72) |
| Pure hypercholesterolemia | | 1.462 (1.118-1.911) | 1.47 (0.721-2.997) |
| Hypertriglyceridemia | | 0.977 (0.675-1.415) | 1.33 (0.628-2.82) |
| Hyperlipidemia | | 0.996 (0.832-1.193) | 1.002 (0.678-1.48) |
| Ischemic heart disease | | 0.939 (0.746-1.183) | 0.99 (0.59-1.66) |
| Other Heart disease | | 0.948 (0.729-1.232) | 1.313 (0.711-2.427) |
| Hypertension | | 1.151 (0.973-1.363) | 1.377 (0.944-2.009) |
| T2DM | | 1.146 (0.936-1.403) | 1.099 (0.687-1.758) |
| Cerebral infarction | | 1.93 (1.003-3.712) | 6.054 (2.028-18.069) |
| Atrial fibrillation | | 1.207 (0.787-1.852) | 0.435 (0.061-3.123) |
| Hypothyroidism | | 1.15 (0.783-1.69) | 1.456 (0.571-3.713) |
| Hyperthyroidism | | 1.095 (0.345-3.469) | 2.139 (0.242-18.92) |
| Depression | | 1.129 (0.916-1.393) | 1.159 (0.695-1.934) |
| TBI | | 1.587 (0.65-3.876) | - |
| Alcohol dependence | | 0.727 (0.555-0.953) | 0.621 (0.272-1.421) |
| Parkinson's disease | | 3.662 (2.088-6.423) | - |
| Generalized anxiety disorder | | 1.273 (0.748-2.166) | - |
| Chronic kidney disease | | 0.562 (0.367-0.862) | 0.576 (0.214-1.549) |

HIV, Human immunodeficiency virus. HBV, Hepatitis B virus. BMI, body mass index. T2DM, type 2 diabetes mellitus. TBI, traumatic brain injury. NRTI, nucleoside reverse transcriptase inhibitor.

**Supplementary Table 8. Morris Water Maze, Mixed effect model results.**

| | Time | | |
|---|---|---|---|
| *Predictors* | *Estimates* | *CI* | *p* |
| (Intercept) | 56.59 | 50.24 – 62.95 | **<0.001** |
| 24-week 5xFAD | 8.42 | 1.81 – 15.02 | **0.013** |
| 36-week 5xFAD – K-9 | -10.84 | -18.48 – -3.20 | **0.006** |
| 36-week 5xFAD - PBS | 9.21 | 1.57 – 16.85 | **0.018** |
| WT | -9 | -16.63 – -1.37 | **0.021** |
| day | -8.37 | -9.71 – -7.02 | **<0.001** |
| **Random Effects** | | | |
| $\sigma^2$ | 278.98 | | |
| $\tau_{00}$ Mouse.ID | 20.24 | | |
| ICC | 0.07 | | |
| N Mouse.ID | 30 | | |
| Observations | 478 | | |
| Marginal $R^2$ / Conditional $R^2$ | 0.345 / 0.389 | | |

*All groups compared to reference level of 13-week 5xFAD mice

**Supplementary Table 9. Morris Water Maze, Contrast estimates**

| | Time | | |
|---|---|---|---|
| *Contrast* | *Estimates* | *CI* | *p* |
| 36-week 5xFAD – K-9 vs. 36-week 5xFAD - PBS | -20.05 | -28.66, -11.44 | **<0.001** |
| 36-week 5xFAD – K-9 vs. 18-week wild-type | -1.842 | -10.44, 6.76 | 0.85 |
